# Multisite Study of Optical Genome Mapping of Retrospective and Prospective Constitutional Disorder Cohorts

**DOI:** 10.1101/2022.12.26.22283900

**Authors:** Ulrich Broeckel, M. Anwar Iqbal, Brynn Levy, Nikhil Sahajpal, Peter L. Nagy, Gunter Scharer, Aaron D. Bossler, Vanessa Rodriguez, Aaron Stence, Cindy Skinner, Steven A Skinner, Ravindra Kolhe, Roger Stevenson

**Affiliations:** Section of Genomic Pediatrics, Department of Pediatrics, Medical College of Wisconsin, Milwaukee, WI, USA; DNA Microarray CGH Laboratory, Department of Pathology, University of Rochester Medical Center, Rochester, NY, USA; Columbia University Medical Center, New York, NY, USA; Greenwood Genetic Center, Greenwood, SC, USA; Praxis Genomics, Atlanta, GA, USA; H. Lee Moffitt Cancer Center, Tampa, FL, USA; University of Iowa Hospitals & Clinics, Iowa City, IA, USA; Department of Pathology, Medical College of Georgia, Augusta University, Augusta, GA, USA; Equanimitas, Greenwood, SC, USA

## Abstract

Several medical societies including the American College of Medical Genetics and Genomics, the American Academy of Neurology, and the Association of Molecular Pathology recommend chromosomal microarray (CMA) as the first-tier test in the genetic work-up for individuals with neurodevelopmental disorders such as developmental delay and intellectual disability, autism spectrum disorder, as well as other disorders suspected to be of genetic etiology. Although CMA has significantly increased the diagnostic yield for these disorders, limitations in the technology preclude detection of certain structural variations in the genome and requires reflexing to other cytogenomic and molecular methods. Optical genome mapping (OGM) is a high-resolution technology that utilizes ultra-high molecular weight DNA, fluorescently labeled at a hexamer motif found throughout the genome, to create a barcode pattern, analogous to G-banded karyotyping, that can detect all classes of structural variations at very high resolution by comparison to a reference genome.

A multisite study, partially published previously, with a total of n=1037 datapoints was conducted and showed 99.6% concordance between OGM and standard-of-care (SOC) testing for completed cases. The current phase of this study included cases from individuals with suspected genetic conditions referred for cytogenomic testing in a prospective postnatal cohort (79 cases with OGM and SOC results) and a retrospective postnatal cohort (262; same criteria). Among these cohorts were an autism spectrum disorder cohort (135) group with negative or uninformative results on previous testing (72). Prospective cases referred for CMA were included in this study as an unbiased comparison, OGM results show 100% concordance with variants of uncertain significance, pathogenic variants, and likely pathogenic variants reported by CMA other SOC and found reportable variants in an additional 10.1% of cases. Among the autism spectrum disorder cohort, OGM found reportable variants in an additional 14.8% of cases. Based on this demonstration of the analytic validity and clinical utility of OGM by this multi-site assessment, and considering clinical diagnostics often require iterative testing for detection and diagnosis in postnatal constitutional disorders, OGM should be considered as a first-tier test for neurodevelopmental disorders and/or suspicion of a genetic disease.

## Introduction

Genomic testing for constitutional disorders, including multiple congenital anomalies and neurodevelopmental disorders (NDDs), is necessary for accurate diagnosis, clinical management, genetic counseling, and family planning. Developmental disabilities affect about 17% of the pediatric population in the United States^1^, and structural variations (SVs) contribute significantly to NDDs and congenital disorders^2^. In 2010, the ACMG published a recommendation for chromosomal microarray (CMA) testing as the first-tier test for these patients, effectively replacing G-banded karyotype as the frontline genome-wide method^3,4^. In subsequent years, other professional organizations followed, largely due to the higher resolution and diagnostic yield of CMA over karyotyping (KT) (15-20% versus 5%, respectively)^5,6^. However, CMA has several technical limitations such as inability to detect balanced rearrangements, repeat expansions, SVs smaller than ≈40 kbp, and the genomic location and orientation of copy number gains^7^. As a result of these limitations, use of additional testing modalities is often necessary. For example, for individuals with NDD, including intellectual disability and autism spectrum disorder, Fragile X Syndrome (FXS) testing is also recommended as an ancillary test^7^.

Since 2019, the ACMG has recommended exome sequencing (ES) for children with NDD^8^, and in 2021 ES or genome sequencing (GS) were recommended for congenital anomalies and intellectual disability^9^ as either the first or second tier test due to an estimated diagnostic yield of ≈35%^10^. However, detection of SVs and repeat expansions and contractions with ES/GS is limited^11^. Benchmarking of short read sequencing has clearly shown these limitations of GS for SV detection^12^, hence cytogenomic methods remain essential in postnatal genomic testing.

Ancillary methods for repeat disorders analysis such as Southern blotting or PCR testing are still required for repeat expansion/contraction analysis, but these methods are time-consuming and have low accuracy (Southern blotting; e.g. for facioscapulohumeral muscular dystrophy (FSHD1)) or have a low sizing range (PCR, NGS; e.g. for FXS)^11^. Taken together, individuals with NDDs or congenital anomalies are likely to have multiple testing methodologies performed in the quest for a diagnosis, and for some, this diagnostic odyssey may take many years, with some never achieving a diagnosis.

Optical genome mapping (OGM) is a next generation cytogenomic technique that enables higher resolution compared to cytogenetic/cytogenomic technologies and comprehensive analysis of the genome for CNVs and SVs including repeat expansions^13^. Using linearized, ultra-high molecular weight (UHMW) DNA molecules (≥150 kbp) labeled at a 6 base-pair sequence motif found every 5 kbp on average throughout the genome, OGM can detect all classes of SVs from 500 bp (insertions, deletions), or >≈30 kbp (inversions and translocations), triploidy, and absence of heterozygosity (AOH) segments (≥25Mbp in Solve 3.7.x)^14-16^. The consensus maps of linearized DNA molecules are compared to a reference genome to find SVs and a separate coverage-based algorithm allows for the detection of additional CNVs. OGM is the only technology that is capable of detecting and sizing large repeat expansions and contractions, genome wide, including sizing of CGG repeat expansions in the *FMR1* gene (FXS) and D4Z4 repeat contractions with terminal haplotype (4qA or 4qB: permissive or benign haplotypes for FSHD1, respectively)^17^. Importantly, the OGM workflow is comparable to other SOC test protocols, and results can be available within five days.

The first phase of this multisite evaluation was used to validate the performance of OGM compared to SOC technologies (CMA, KT, FISH, Southern blotting, and PCR), in a cohort of 404 datapoints^18^ (accepted for publication in the Journal of Molecular Diagnostics, in press, 2022). In the current study, 560 unique case analyses were used to evaluate the technical and clinical utility of additional undiagnosed postnatal cohorts. Other primary endpoints include performance of OGM in a prospective cohort against SOC (n=79); performance of OGM in an autism spectrum cohort (n=135) with negative or uninformative results by SOC (n=72); and family analysis to demonstrate inheritance of CNVs or SVs (41 families). This study was performed to assess the performance of OGM in a multi-centric model (multisite, multi-operator, multi-instrument) in a cost-effective manner for challenging diagnosed and undiagnosed rare diseases. As available, duo/trio studies were performed to establish the mode of inheritance and pathogenicity or diseases segregation within the family. The results demonstrate that OGM can detect relevant genomic aberrations that mitigate the need for numerous testing platforms and time-consuming wet lab work, potentially improving patient care by reducing the associated time and costs for the diagnostic odyssey.

## Methods

### Case Cohort Design, and Participating Laboratories and Reviewers

Blood or cell samples corresponding to known or suspected postnatal disorder cases were recruited for this study. Samples were given anonymous aliases used in this study (i.e.: PUBKH-xxxxx). Data from one or several of the standard of care (SOC) methods, including karyotyping, FISH, CMA, Southern Blotting, and/or PCR were collected for the final unblinded analysis to compare the performance against OGM. A companion effort^18^ includes datasets published previously, for which totals of the larger effort are summarized in Table 1. For the scope of this publication, 560 unique postnatal datasets are reported with novel analyses.

**Table 1.** Progress report and technical concordance summary of postnatal data collection and analysis between Iqbal et al. 2022 and this preprint.

|  |  |
| --- | --- |
| **Total datasets collected** | 1037 |
| **Datasets redundant with others as replicates** | 180 |
| **Datasets with variants not fitting design, mixups, or with unclear or incomplete classification for this preprint** | 108 |
| **Total cases assessed for:** | **Technical Concordance (unblinded)** |
|  | 749 |
| **Concordant** | 739 / 749 (98.7%) |
| **Partially Concordant** | 7 / 749 (0.9%) |
| **Discordant** | 3 / 749 (0.4%) |

Nine U.S. based laboratories contributed to the study for sample recruitment, data collection, variant analysis (**Figure S1)**. Upon bioinformatic analysis and confirmation of technical concordance to SOC by one unblinded analyst, individuals from nine institutions across the Unites States contributed to variant curation and analyst review. Geneticists and clinical laboratory directors performed the director level review to finalize all cases.

The case cohorts were defined based on the clinical indications provided with the sample. Neurodevelopmental problems including autism and/or autism spectrum disorder (ASD) and intellectual disability/development disorder (ID/DD) constituted a majority of the indications for testing, often in combination with other indications/ categories (**Figure 1a**). Other indication categories included attention deficit hyperactivity disorder/oppositional defiant disorder (ADHD/ODD) and seizures.

**Figure 1.**
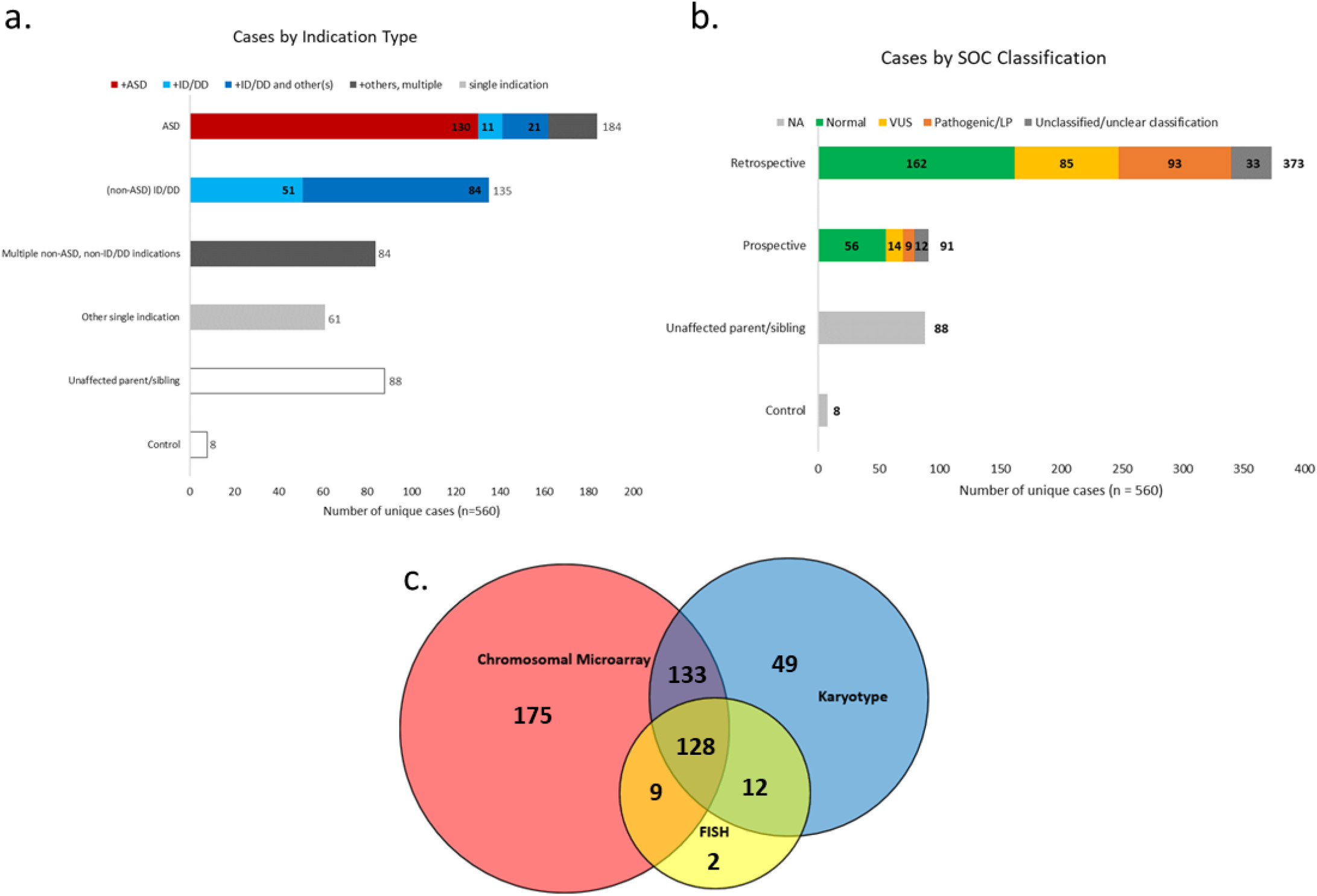
**a**. Unique postnatal cases (n=560) represented by clinical indication grouping. **b**. Schematic of unique postnatal cases represented across cohorts characterized, striated by SOC classification grouping. Autism spectrum disorder (ASD) and Intellectual disability / developmental delay (ID/DD) shown as priority in hierarchy **c**. Standard of care (SOC) cytogenomic methods summary used in applicable (n=508) postnatal cases. 52 cases had method(s) outside this modality, i.e. targeted XON array in unaffected parents

Further, major cohort categories were defined for these postnatal cases as follows: retrospective (n=373), prospective (n=91), family studies (n=130), and healthy controls (n=8) (**Figure 1b**). 39 cases fit both the retrospective and family cohort—for example, a child with a still-undiagnosed genetic disease after undergoing diagnostic odyssey, with parent(s) enrolled in the study, fits both cohorts. Similarly, three cases fit both the prospective and family cohorts.

This IRB approved study included consent provided by individuals with newly collected samples or waived authorization for use of de-identified samples. All protected health information (PHI) was removed, and data were anonymized (coded and double-blinded) before accessioning for the study.

### OGM Data Collection and Analysis

The workflow from tissue sample through blinded case analysis is summarized in **Figure S2**. Frozen aliquots of whole blood and/or lymphoblastoid cell lines in cryoprotectant served as the sample tissues. Samples were collected and shipped on dry ice to a central laboratory where they were accessioned with common study identifiers. Samples were then assigned and shipped to a participating OGM data collection site with a Saphyr System(s), and DNA isolated for downstream analysis by the receiving site.

Frozen sample vials were thawed in 37°C water baths and counted for number of cells. Thawed, mixed liquid volumes representing 1.5 million viable cells per sample were transferred to Protein Lo-Bind microfuge tubes for centrifugation. With the target number of cells pelleted, crude supernatant was removed and cells washed in stabilizing buffer. Washed cell suspensions were enzymatically digested and lysed, and isopropanol used to precipitate UHMW DNA in the presence of a nanobind disk. Long DNA strands bound on disks were further washed, then transferred to clean tubes and eluted in buffer.

750ng of solubilized UHMW DNA was labeled enzymatically, conjugating fluorophores to the target 6-mer *CTTAAG* without cleaving the phosphate backbone. Long, labeled DNA strands were then counterstained with an intercalating dye, homogenized in buffer to promote flow through a nanochannel device, then loaded into flowcells of Saphyr G2.3 chips. Chips were run in Saphyr Systems to a target throughput of 800 Gbp per flowcell. Completed datasets were then assessed for analytical quality control—targeting 160× effective coverage of GRCh38, with ≥70% of molecules ≥150 kbp aligning (“map rate”) and at an N50 of ≥230 kbp.

Completed datasets were uploaded to a central server running Bionano Access versions 1.7 or 1.7.1. (hereafter, v1.7.x) for analysis with Bionano Solve 3.7 or 3.7.1. (hereafter, v3.7.x). Study datasets were managed across participating laboratories, analysts, and directors—within shared directories. GRCh38 was selected as reference, and bioinformatic analyses were launched from Access to Bionano Solve pipeline software as described in its documentation^19^ and subsequent sections.

### Bioinformatic analyses

#### EnFocus™ Repeat Analyses

Bionano Solve 3.7.x software supports two *EnFocus*^*TM*^ pipelines, each targeting a region implicated in a repeat array disorder— FSHD and FXS. The EnFocus™ pipelines report a stable region assembly analysis as a useful post-analytic QC measure. An EnFocus™ Fragile X Analysis^20^ was run for every case to detect for *FMR1* full expansions. For cases with known or suspected indications of FSHD1, an EnFocus™ FSHD Analysis was conducted.

### De Novo Assembly

The Bionano Solve 3.7.x De Novo Assembly for genome-wide bioinformatic analysis was used. Briefly, this pipeline uses pairwise alignment and assembly of labeled molecules to generate haplotype-refined maps with consensus label positions and inter-label distances. These maps were aligned to the GRCh38 reference *in silico* digested with the *CTTAAG* label motif, and a subsequent SV calling algorithm run to catalog discrepancies in sample × reference alignment as structural variants; in terms of size, zygosity, and variant allele fraction (VAF), among other properties. SV calls are finally annotated based on positions in the GRCh38 reference and queried against a database of 179 population control assemblies among other annotations^21^.

A built-in absence of heterozygosity (AOH) detection tool uses variant zygosities and their positional information to identify likely AOH segments. This process analyzes the distribution of homozygous and heterozygous variants spatially along reference chromosomes and identifies segments (≥25Mbp) with higher-than-expected homozygous variants likely to represent AOH based on distributions in control samples. Separately, the analysis generates a copy number profile to call copy number gains and losses analogous to microarray. Molecules are aligned to the GRCh38 reference to create a depth of coverage profile; which is then normalized based on label motif-specific controls, putative outliers are segmented, then scaled against a baseline defined at CN=2 in autosomes (X and Y have a sex chromosome-specific baseline). CNV calls are generated and similarly annotated with positional information from the original reference. Entire chromosomal aneuploidies are likewise defined in the CNV algorithm.

### Post-Analytical Variant Curation and Classification

An analyst (designee) was assigned to remain unblinded to the SOC finding(s) available for each case, for this study. On completion of each case’s bioinformatic analysis, this individual checked the OGM results for technical concordance with SOC finding(s). Cases not technically concordant with SOC were investigated further; those meeting technical concordance proceeded through an establish standard operating procedures for review and analysis of the OGM data with blinded analysts and directors.

To establish consistency and harmonization of the OGM data analysis across multiple sites and investigators, a filtering and curation protocol was devised for the blinded review process of each case using Bionano Access v1.7.x. The De Novo Assembly workflow above typically generates 5000-6500 variant calls per sample aligned to GRCh38, which pass basic quality criteria^18-19^. The filtering workflow reduces the burden for SV analysis to a manageable number that are more likely to be clinically relevant—typically 25-60 variants per case. It proceeds by the following steps, shown graphically in Figure S2:

- The analyst first prioritizes large-scale abnormalities: aneuploidies, AOH segments, and evidence of triploidy. Presence or absence of these abnormalities is recorded by the analyst in the case summary notes, along with an ACMG classification if one is applicable^22^. The inferred copy number baseline of sex chromosomes serves as sex determination in OGM analysis and is likewise recorded in this step along with consistency with the case’s sex indication at the time of sample collection.
- The analyst then manually examines three regions of GRCh38, in which automated analysis could show false negative calls of known microdeletion/microduplication syndromes. These regions are chr16:21,935,203-22,455,963 (recurrent 16p12.1 microdeletion), chr16:29,595,531-30,188,534 (16p11.2 microduplication syndrome), and chrX:154,396,222-154,653,579 (Xq28 microduplication). Any variant reflecting these findings is manually added to the curated list.
- Genome-wide interstitial variants are filtered by size (≥1500bp), rarity in the population controls database (≤1%) and overlap with GRCh38 known canonical genes within 3 kbp breakpoint precision. Masking of SV/CNV calls is removed as a filter. Variants meeting these criteria are added to a curated list within Bionano Access, for subsequent review and classification.
- Any Inter-Translocation or Intra-Fusion calls meeting baseline quality criteria are added to the curated list, regardless of gene overlap.
- At the analyst’s discretion, variants not meeting the criteria above may be added manually to the curated variant list based on gene/disease association for final Director review. This flexibility allowed for the most comprehensive analysis of all “suspicious” SVs in a minority of cases.

The analysts then proceeded to classify variants in the curated list using ACMG criteria adapted for OGM data^22^. Suspected technical artifacts were assigned an “Unclassified” designation, with explanation given in variant notes section of each case.

Completed analyst reviews were sent to blinded directors for director-level classification and sign-out. Directors proceeded through classified variants in the curated list, upholding and/or revising analyst classification as necessary. Upon completing review of the curated variant list, directors affirmed or modified case summary notes regarding large-scale abnormalities and documented a completed case review with director’s notes on large-scale events and adjudicated variant classifications.

### Family-based OGM Analysis

Members (a total of 117 individuals) from 41 families underwent additional analysis to determine inheritance pattern of clinically significant variants identified in the probands. Bionano Access and Solve support Dual and Trio analyses, using the Variant Annotation Pipeline^21^. Briefly, this workflow accepts two or three singleton assemblies with SV calls to a common reference as input. One assembly is designated as the proband, whose SV calls are queried against those in one matched control (dual) or two parent control (trio) datasets. SVs in the proband are sought through alignment of assembled maps and raw molecules of matched control(s). If present, the proband SV is annotated as found in matched control(s).

41 cases were enrolled as probands of which seven were run in duo analysis (only one available parent), 33 were run in trio analysis (both parents), and one run in quad analysis (both parents and one sibling). Proband SVs found in a matched control/parent were considered inherited; those absent in both parent assemblies/molecules are assessed to be *de novo* variants.

## Results

### Overall concordance

OGM data have been collected for 1037 datapoints to date for postnatal analysis, including 404 postnatal datasets published in a companion paper to this effort^18^ After removing 180 datasets deemed redundant (and replicates), 820 unique cases are represented; 760 of which have completed OGM classification at time of this pre-print using the methods outlined above. Three cases were FXS indications with inconclusive OGM EnFocus^™^ Fragile X results, and an additional eight are awaiting SOC results, so a total of 749 was used to calculate technical concordance.

Overall, 746 / 749 (99.6%) datapoints were fully or partially technically concordant with SOC results: 739 datapoints were fully concordant with SOC results (98.7% full technical concordance) with seven datapoints (0.9%) partially concordant, and three datapoints (0.4%) discordant (Table 1).

### Prospective case analysis

This study included 91 prospective cases from two sites. SOC results were not known for these cases at the time of sample collection for OGM. All samples yielded quality OGM data for analysis and interpretation. There were 46 female and 45 male cases in this cohort, and the age at specimen collection ranged from 2 days to 49 years. 38 cases had a neurodevelopmental phenotype including ASD, ID/DD, ADHD/ODD, and/or seizures; while 21 of these cases also had other clinical features including multiple congenital anomalies (MCA), dysmorphic features, skeletal anomalies, and hypotonia. Overall, 23 cases had a clinical indication of MCA. In addition, there were 25 adult cases referred for testing of which 14 cases had history of recurrent pregnancy losses while the other 11 cases had a family history of a child with a previously detected finding and/or a child with clinical phenotype.

Technical concordance has been completed for 89 cases, 100% of which were fully technically concordant. Review and classification of all the OGM data is complete for 79 (of 89) cases. We define reportable yield in this study as any pathogenic or likely pathogenic finding; or VUS finding that was reported (in SOC), or sufficiently unique and compelling (in OGM) to pursue confirmatory testing of functional impact in context of the case’s clinical indication. Of the 79 prospective cases with both SOC and OGM completed, SOC found nine (11.4%) cases with at least one pathogenic or likely pathogenic classification, and ten (12.7%) results with a VUS as the highest reported classification— for an SOC total reportable yield of 24.1%, of which all were also found with OGM. Among prospective cases without a VUS, likely pathogenic, or pathogenic finding in SOC—OGM found compelling VUSs in seven additional cases (Table S2), and a likely pathogenic finding from OGM was detected in one case which did not have classified results on previous testing. This gain of eight cases represents an increase in reportable yield of 10.1% over standard of care, with total combined (SOC and OGM) reportable yield among the prospective cases at 34.2%.

OGM provides high resolution SV refinement: A particular case (PUBKH-00869) demonstrated the benefit of OGM to help resolve structure. The terminal loss of chromosome 9 and contiguous proximal copy number gain observed in the SOC was resolved by the OGM results showing a foldback mechanism to rescue the terminal loss of 9p. Subsequent karyotyping confirmed the OGM finding. A second case with a potentially significant incidental finding (PUBKH-01036) is that of an adult patient with an indication of recurrent pregnancy losses. OGM revealed an insertion in intron 1 of the *C9orf72* gene (confirmation pending), which is associated with frontotemporal dementia and/or amyotrophic lateral sclerosis 1. This autosomal dominant condition is caused by heterozygous hexanucleotide repeat expansion (GGGGCC) in intron 1 of the *C9orf72* gene. Although this finding does not explain the recurrent pregnancy losses, upon agreement with board-certified Director review, this incidental finding may have significant clinical implications. Details of the results are provided in Table S1.

### Autism spectrum disorder (ASD) cohort

The study included 184 cases with autism or ASD given in the clinical indication. From the 184 ASD total— 130 had indications solely consisting of ASD, while 32 also referenced ID/DD. The remaining 22 cases indicating ASD referenced one or more other indication categories (Figure 1A). Those individuals with complex clinical symptoms were more likely to have an abnormal SOC result: Of the 130 cases with only ASD as the indication, 44 cases had an SOC result reporting a VUS, likely pathogenic, or pathogenic finding; giving a reportable yield of 33.8% (44/130). Of those 54 with additional clinical phenotypes, 35 such cases had a reported SOC result for a reportable yield of 64.8% (35/54).

Review and classification of both the SOC and OGM has been completed for 135 of the 184 ASD cases. Of these 135, SOC found 22 (16.3%) of cases with at least one pathogenic or likely pathogenic classification, and 41 (30.4%) results with a variant of uncertain significance (VUS) as the highest reported classification— for an SOC total reportable yield of 46.7%. Among ASD cases without a VUS, likely pathogenic, or pathogenic finding in SOC—OGM found compelling VUSs in 18 additional cases (Table S3), and pathogenic/likely pathogenic findings in two additional cases. This gain of 20 cases represents an increase in reportable yield of 14.8% by OGM, with total combined (SOC and OGM) reportable yield among the autism spectrum disorder cases at 61.5%.

Four ASD cases had additional OGM findings that were classified as pathogenic or likely pathogenic. Two had normal SOC results from CMA testing, and two reported a VUS. Currently, one of the SOC normal cases (PUBKH-00847, Figure 2c) has already been confirmed—a multi-exonic deletion within *AP1G1*, associated with recently described autosomal dominant Usmani-Riazuddin syndrome (OMIM 619467)^23^. Usmani-Riazuddin syndrome is characterized by mild to severe intellectual disability, epilepsy, and developmental delay.

**Figure 2.**
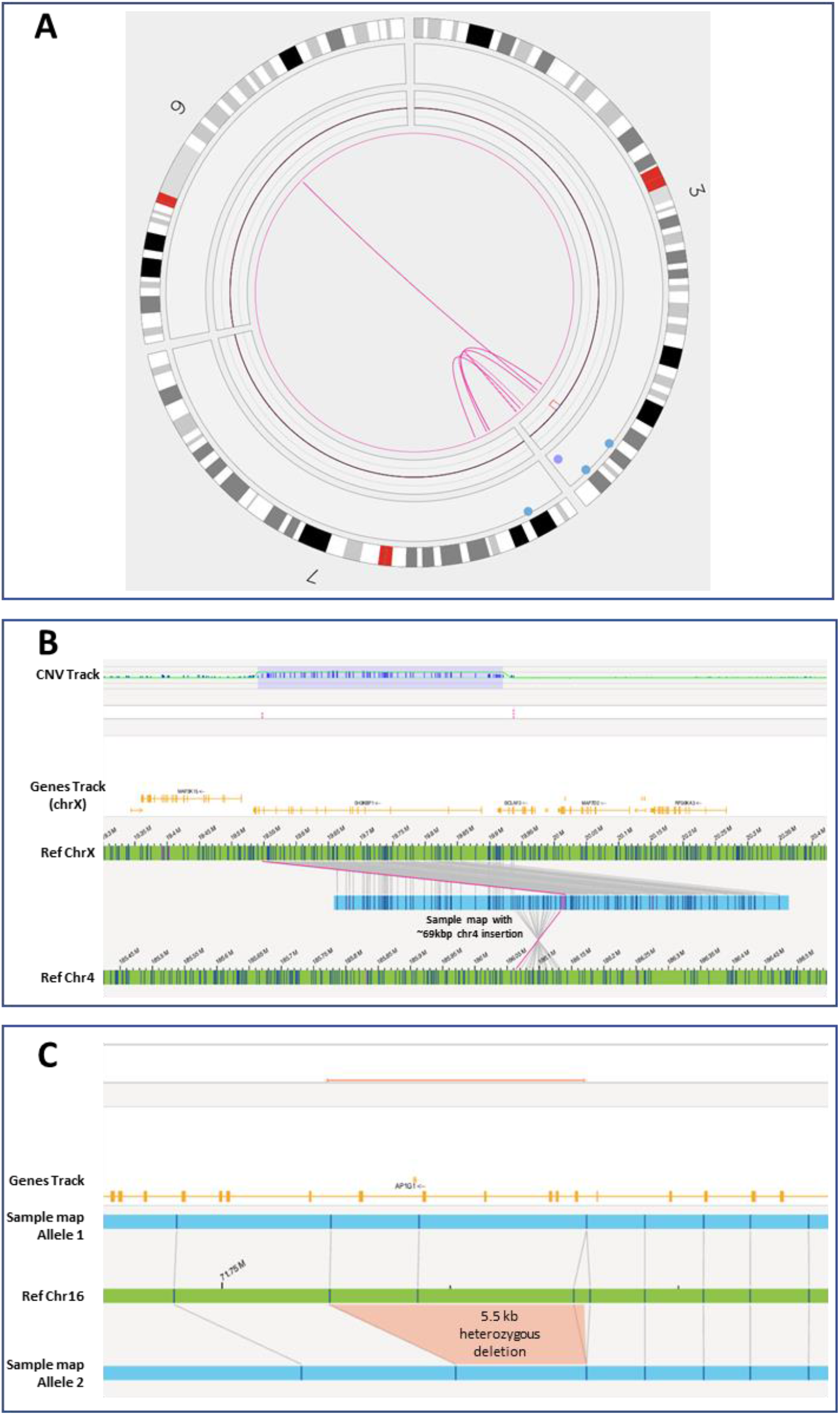
Novel findings by OGM, **in a**. an unclassified retrospective case, PUBKH-00711, with indication of anophthalmia, hydrocephalus, and development delay. The case underwent karyotype, which reported a t(3;7)(q29;p22.3) but did not provide a classification for the translocation or note any impact to copy number. OGM revealed a more complex rearrangement involving three chromosomes, involving several translocation breakpoints and intrachromosomal foldback junctions. Included is a 2.78 Mbp copy number loss over SOX2 gene, which is related to eye and optic nerve problems with reported cases of anophthalmia and microphthalmia. **b**. a prospective case, PUBKH-00813. OGM revealed the Xp22.12 duplication structure, with a 69 kbp duplication from chr4 (186,063,759-186,132,836) inserted in between the 390 kbp tandemly duplicated segment (chrX:19,547,445-19,937,089). The inserted segment from chr4 contains the entire TLR3 gene and exon 1 of FAM149A, and its orientation and position supports a potential fusion with BCLAF3 on chrX. **c**. a retrospective, autism spectrum disorder case with normal SOC reported (PUBKH-00847). A heterozygous 5.5 kbp deletion in AP1G1, size and boundary of which suggests impact to exons 11-12 or 12-13. Subsequent analysis with qPCR confirmed deletion of exons 11 and 12. AP1G1 is related to autosomal dominant Usmani-Riazuddin syndrome (https://omim.org/entry/619467).

**Figure 3.**
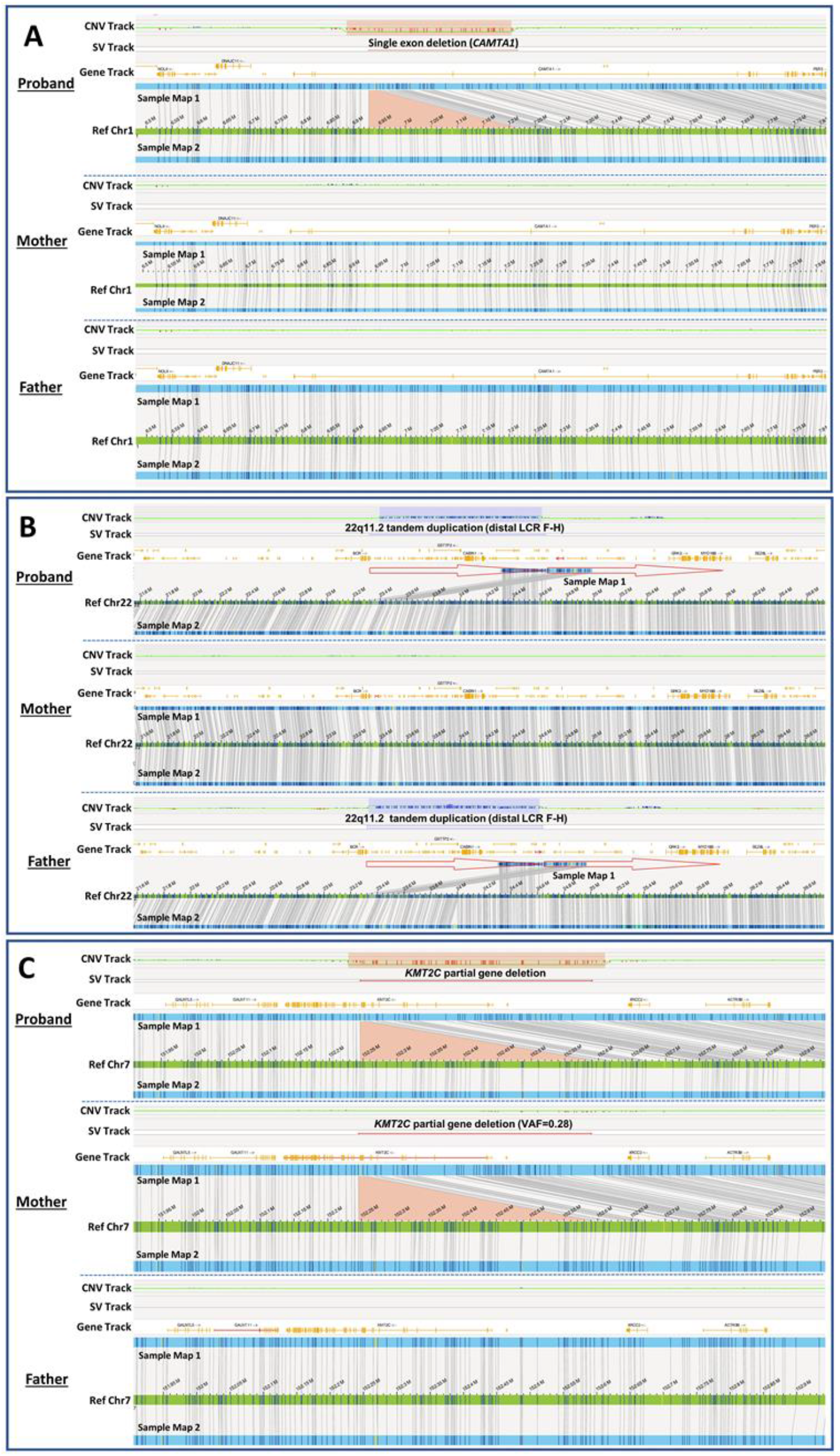
Findings from family-based cohort using dual/trio-based analyses **a**. A de novo 272 kbp heterozygous deletion identified in the proband (PUBKH-00141) was absent in the parents. The deletion is highlighted in CNV (pink shade) and SV track (red line). This deletion affects exon 4 of CAMTA1 (NM_015215.4) and likely results in frameshift and premature termination. **b**. A 1.3 Mbp heterozygous 22q11.2 distal LCR F-H tandem duplications were found in the proband (PUBKH-00135) and the father. The duplication is highlighted in CNV track (purple shade) and SV track (purple line). Tandem duplications are illustrated as red arrows. **c**. A 338 kbp heterozygous deletion potentially affecting exon 1-14 of KMT2C (NM_170606.3) was identified in the proband (PUBKH-00118) and the mother. The deletion is highlighted in CNV (orange shade) and SV track (red line). However, the deletion found in the mother had a variant allele fraction (VAF) of 0.28, suggesting possibility of mosaicism.

### Family-based analysis

Members of 41 families underwent family-based analysis to determine inheritance pattern of clinically significant variants identified in the probands. Details of the results are provided in Table S4. 23 cases had d*e novo* variants identified in the probands, and the majority (20) of *de novo* variants were classified as pathogenic or likely pathogenic (P/LP). One example is case PUBKH-00141 (Figure 4A), a young male with speech delays, hypotonia, ASD, learning difficulties, ADHD, multiple episodes of febrile seizures, and aggressive behaviors. OGM identified a *de novo* 272 kbp heterozygous deletion affecting exon 4 of *CAMTA1* associated with cerebellar dysfunction with variable cognitive and behavioral abnormalities (MIM: 614756). This deletion is similar to a previously known pathogenic 81 kbp deletion of exon 4 resulted in frameshift and premature termination^24^. There were three *de novo* variants classified as VUS including a 300 kbp 16p13.3 deletion (chr16:2,315,760-2,615,921, GRCh38), a 685 kbp 20p13 duplication (chr20:2,456,048-3,140,786, GRCh38), and a 7.8 Mbp 8q24.2 deletion (chr8:128,253,843-136,061,268, GRCh38). Classification of 8q24.2 deletion in case PUBKH-00819 was later changed to likely pathogenic because of identification of *de novo* origin.

13 probands carried clinically significant variants that were inherited from one of their parents. Inherited variants in the proband that were classified as P/LP include a 1q21.1 duplication (BP3-BP4, distal), a 16p13.11 duplication (BP1-BP3), a 22q11.2 duplication (LCR F-H, distal), an 8p23.1 duplication, a 16p12.2 deletion (proximal), a 10q11.2 deletion (LCR C-D), and a recombinant X chromosome with Xpter duplication and Xqter deletion. Variants in these regions were known to have lower penetrance or variable expressivities, which explains why parents of probands who also carry these variants may be asymptomatic or have milder symptoms^25-29^. For example, OGM identified a 22q11.2 duplication (LCR F-H, distal) in a young male (PUBKH-00135) who had global delays and ASD, and his apparently healthy father also carries the same duplication (Figure 4B). In another case, a young female (PUBKH-00067) with congenital hypotonia and delayed milestones, inherited a recombinant X chromosome with Xpter duplication (ogm[GRCh38] Xp22.33p22.2(2720069_10542462)x3) and Xqter deletion (ogm[GRCh38] Xq28(154591265_156025612)x1) from her mother who had lupus and rheumatoid arthritis. Terminal X chromosome duplication (Xpter) and deletion (Xqter) had been reported to have variable expressivity and reduced penetrance as well as skewed or complete X-inactivation in females^30-32^.

Another informative case (PUBKH-00118) was of a young male with clumsiness, drooling, sensory disorder, speech articulation disorder, developmental coordination disorder, facial weakness, and hypotonia. OGM identified a multi-exon deletion of *KMT2C* which is associated with Kleefstra syndrome type 2 (MIM: 617768). However, this deletion was also found in his healthy mother (Figure 4C). When we further examined variant allele fraction (VAF), *KMT2C* deletion in the mother had a lower VAF of 0.28, suggesting potential mosaicism. Confirmatory testing for the targeted *KMT2C* deletion was performed by CMA and revealed low-level mosaicism. Although mosaicism in apparently unaffected parents had been previously reported in Kleefstra syndrome type 1 (MIM: 610253) caused by mutations in *EHMT1* (Ref PMID: 29416845), mosaic *KMT2C* pathogenic variants have only been reported in patients^33,34^ but not in unaffected parents or healthy individuals to our knowledge.

### Retrospective cohort analysis

Among the 373 retrospective cases, 340 were assessed for technical concordance, with full or partial technical concordance achieved in 337 (99.1%) cases. Additional pathogenic or likely pathogenic findings from OGM were detected in six cases, two of which had normal results on previous SOC testing. One such case was PUBKH-00711 (Figure 2a)—a teenaged male with anophthalmia, hydrocephalus, undergrowth, and development delay. The case was referred for karyotype, which reported an t(3;7)(q29;p22.3). The original SOC did not reference any CNV and did not provide a classification for the translocation finding. OGM revealed a more complex rearrangement involving 3q29, 7p21.1, and 9q21.2; involving six separate fusions, intrachromosomal foldback junctions, and a 2.78 Mbp copy number loss involving the *SOX2* gene (Figure 2a). *SOX2* disruption is associated with ocular and optic nerve abnormalities including anophthalmia and microphthalmia.

In a case of a young female with ASD and self-injurious behaviors (case PUBKH-00412) previous CMA found two copy number changes, a pathogenic chr7 gain and a chr9 loss of uncertain significance (arr[GRCh37] 7p22.3p21.1(43,376_19,520,619)x3, 9p24.3(203,861_1,539,085)x1). Given that both are terminal, the reporting lab suggested a possible unbalanced translocation. Analysis with OGM revealed the complete structure which was an unbalanced translocation with intergenic breakpoints (ogm[GRCh38] t(7;9)(p21.1;p24.3)(19,469,714;1,550,014)).

## Discussion

One of the most compelling benefits of OGM in postnatal constitutional clinical research is the ability to detect both balanced and unbalanced SVs at very high resolution – down to 500 bp for the SV algorithm and to 30 kbp for the CNV algorithm. With this level of resolution and the detection of balanced and/or cryptic rearrangements, the diagnostic yield for OGM in the constitutional setting would be expected to be higher than CMA. Indeed, in the prospective arm of this study, reportable yield was higher utilizing OGM (34.2%) compared to just that for SOC methods (CMA, karyotype, and FISH) combined at 24.1%. For the autism spectrum disorder cohort, reportable yield utilizing OGM was 60.7% versus 46.7% using only current SOC methods.

The increase in reportable yield by OGM was directly attributable to genomic aberrations that were not discovered using SOC methods. Much of this increase stems from small (single to tens of kbp) intragenic deletions or insertions classified as VUSs, with potential genomic impact too small to be detected by CMA (Tables S2 and S3). One example is that OGM detected a multi-exonic, intragenic deletion of AP1G1 (exons 11 and 12; NM_001030007.2) in case PUBKH-00847, which was negative by SOC (Figure 2c). This intragenic deletion was subsequently confirmed by qPCR assay (unpublished data) thereby highlighting the higher sensitivity of OGM. This case illustrates the importance of gene disruption discovery that allows for the scientific community to advance and also impact the diagnostic odyssey journey of patients and families.

Because of the technical limitations of current cytogenomic testing methods, many patients remain undiagnosed, and the search for answers, known as the diagnostic odyssey, can take years, with many never achieving a diagnosis. Utilizing OGM as a tier 1 approach along with current SOC tests and increasing resolution will significantly increase diagnostic yield and decrease the time of diagnostic odyssey in many cases. Multiple studies have shown that OGM increases the diagnostic yield in SOC negative rare disease cases by at least 10%^35-38^.

When CMA results are not informative, exome and genome sequencing (ES/GS) have been added to the clinical testing options for many individuals affected with a genetic disease^39^. However, several challenges regarding the high cost of instrumentation, bioinformatics core/expertise, and lack of streamlined analysis and reporting tools limit the implementation and reporting of ES/GS in a standardized manner^39-40^. Multiple studies have also revealed the utility of OGM complementing sequencing technologies in resolving rare undiagnosed diseases such as atypical teratoid rhabdoid tumors (ATRT) and inherited retinal diseases^37,38^. For the majority of the undiagnosed rare diseases, the time and costs associated with the diagnostic journey can be staggering on both the families and healthcare systems^41-43^. In fact, the ACMG Policy Statement specifically comments on the clinical utility of attaining a definitive disease on patient management, therapeutic implications, positive impact on patients and their relatives’ psychological status, and overall benefit to the health-care systems^43^.

Multiple studies published by one of the senior authors of this study demonstrate that OGM allows for a streamlined workflow and consolidation of multiple SOC methods for the manner^15,44,45^. By reducing the number of cytogenomic/molecular tests to a single platform with an estimated increase in diagnostic yield by 10-15% (over SOC), one may expect overall healthcare savings and faster time to results thereby ending the diagnostic odyssey of these patients and families.

For most US-based labs, how, whether, and to what extent a clinical test is reimbursed has been a major barrier to adoption^46^. To address this important barrier, two of the senior authors (from Augusta University and Praxis Genomics) on this study have gone through the process of LDT (laboratory developed test) validation and implementation of OGM and obtaining a proprietary laboratory analyte (PLA) code from the American Medical Association (AMA) for CPT™ coding and pricing determination on the Clinical Laboratory Fee Schedule (CLFS) from the Centers for Medicaid and Medicare Services (CMS) (constitutional PLA codes: 0260U, 0264U, 0265U, and oncology PLA codes: 0299U, 0300U, 0331U). Not only is the increase in diagnostic yield by OGM demonstrated in the constitutional setting, but similar increases have been reported by multiple investigators in the heme-onc setting^45,47,48^.

Another major challenge for any existing and new technology to be deployed for clinical testing is impacted by the shortage of technical medical laboratory staff, which is a growing concern and has been well documented by multiple professional societies such as American Society of Clinical Pathology, American Society of Clinical Laboratory Science^49,50^. This challenge has become acute during and after the COVID-19 pandemic affecting turnaround time and workflow efficiencies. Multiple reports demonstrate that OGM can be seamlessly implemented in a routine lab workflow without the need of specialized training for SV detection (as compared to legacy methods) and the end-to-end solution of this assay including the software for data analysis allows for the uniform assessment and interpretation of disease associated structural variants^15,44,45,47,48^. As shown earlier, board certified laboratory directors, geneticists or pathologists performed the final director review in this study thereby demonstrating the effective value of OGM to be used in the detection of structural variants potentially causative of rare diseases^18^.

## Conclusion

This study represents the second phase of a large multisite study on OGMs utility and performance for the detection of all classes of structural variations associated with rare diseases. The data and results highlight OGM as a single platform that can detect all classes of structural variants in the genome (aneuploidies, triploidy, translocations, inversions, microdeletions, microduplications, nucleotide repeat expansions or contractions, and AOH). Numerous cases enrolled in this study required multiple test results from KT, FISH, CMA and PCR and OGM demonstrates its utility not only as a simple and streamlined workflow, but also saving significant healthcare dollars and time. Notably, OGM data analysis was performed by a small team which demonstrates the utility of a consolidated workforce of particular importance for staff shortages or under-resourced laboratories. Lastly, the utility of all the investigators using the same software and analysis pipeline contributes to reducing the time from sample collection to reporting results and also standardization of SV reporting across multiple sites. This large multi-site study is in line with other reports recommending that OGM should be considered as a first-tier test for constitutional postnatal genetic disorders.

## Data Availability

OGM datasets are deidentified, stored, and available by request to the corresponding authors. Details of variant calls are provided in the supplementary tables.

## IRB and disclosures

IRB-20212956 (wcg IRB) – Bionano Genomics Inc., San Diego, CA, USA

IRB-STU#: 1814013– Medical College of Wisconsin, Milwaukee, WI, USA

IRB-STUDY00007572: OGM-Postnatal Study – University of Rochester Medical Center, Rochester, NY, USA

IRB-AAAT9083 – Columbia University Irving Medical Center, New York, NY, USA

IRB-Pro00085001 (Self Regional Healthcare) – Greenwood Genetic Center, Greenwood, SC, USA

IRB-A-#00000150 (HAC IRB # 611298) - Medical College of Georgia, Augusta University, Augusta, GA, USA

RES is a consultant of Bionano Genomics. RK has received honoraria, and/or travel funding, and/or research support from Illumina, Asuragen, QIAGEN, Perkin Elmer Inc, Bionano Genomics, Agena, Agendia, PGDx, Thermo Fisher Scientific, Cepheid, and BMS. All other authors have no relevant disclosures.

## Acknowledgements

The authors wish to thank professional consultants Erica Andersen, Jian Zhao, Bo Hong, and Makenzie Fulmer for support with analysis of cases and preliminary classification of variants. The support to (MAI) from Department of Pathology & Laboratory Medicine, University of Rochester Medical Center is also acknowledged. We also wish to acknowledge the Bionano clinical affairs team: Alka Chaubey, Alex Hastie, Rachel Burnside, Mike Gallagher, James Yu, Ben Clifford, Vruti Mehta, Andy Pang, Jennifer Hauenstein, Alex Chitsazan, Joey Estabrook, Kelsea Chang, Shuk Shukor, and Alberto Reyes for support in training and data management. Finally, we wish to thank all of the patients and families who were willing to contribute to this research study.

## Supplementary material

Table S1: Summary info of 560 unique postnatal cases with novel analyses

Table S2: Prospective cohort compelling OGM VUS’s Table

Table S3: Autism spectrum disorder (ASD) cohort compelling OGM VUS’s Table

Table S4: Family-based OGM Analysis

Figure S1: U.S. map of institutions participating in the multi-site study

Figure S2: Process schematic of blinded sample preparation through director’s review of variants

---

**Figure S1.**
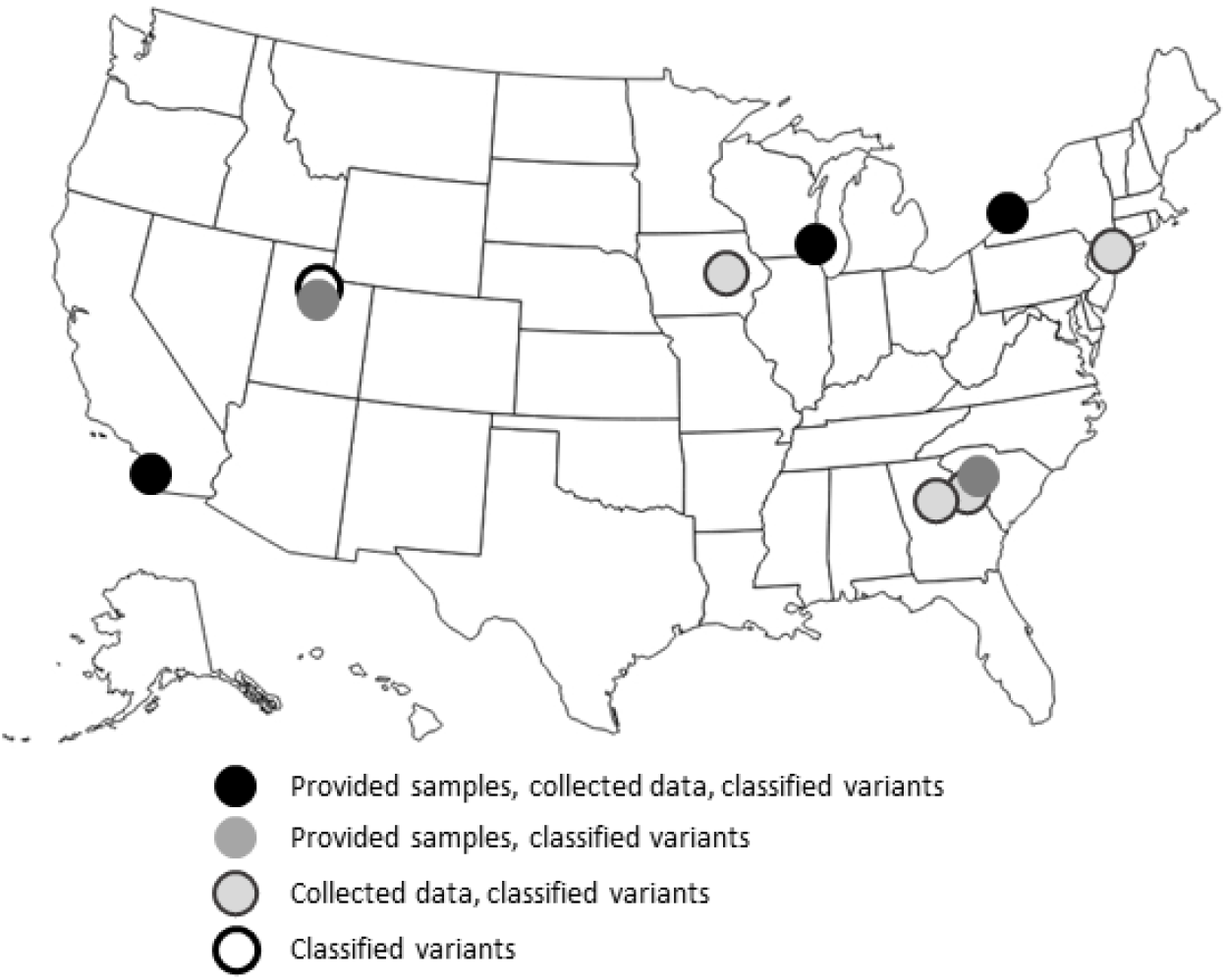
U.S. map of institutions participating in the multi-site study

**Figure S2.**
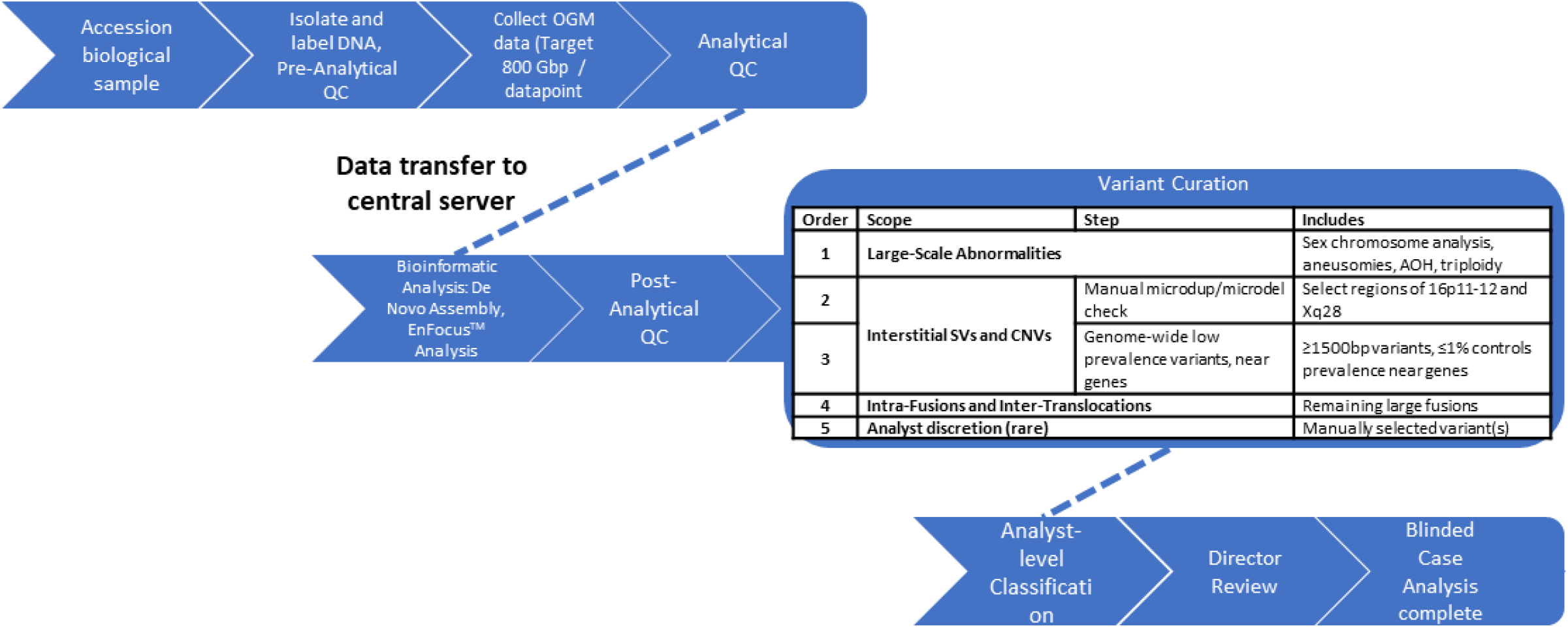
Blinded process schematic of biological sample acquisition through variant analysis, for genome-wide variants assessed with the De Novo Assembly pipeline. A minority of suspected Fragile X and FSHD1 cases were assessed using a targeted EnFocus™ Analysis, and did not proceed to De Novo Assembly and genome-wide curation. (Top row) – Pre-Analytical and Analytical progression of steps, from sample accession through preparation and data collection at laboratory sites. (Middle row) – Centralized bioinformatic analysis through QC and variant curation. (Bottom row) – Variant classification at analyst through director level to case analysis completion.

